# Timeline from receipt to online publication of COVID-19 original research articles

**DOI:** 10.1101/2020.06.22.20137653

**Authors:** Amr F. Barakat, Mohamed Shokr, Joseph Ibrahim, John Mandrola, Islam Y. Elgendy

**Affiliations:** Heart and Vascular Institute, University of Pittsburgh Medical Center, Pittsburgh, PA; Division of Cardiovascular Medicine, Department of Medicine, Wayne State University, Detroit, MI, USA; Department of Internal Medicine, University of Pittsburgh Medical Center, Pittsburgh, PA; Baptist Health Louisville, Louisville, KY; Division of Cardiology, Massachusetts General Hospital and Harvard Medical School, Boston, MA

**Keywords:** COVID-19, medical journals, original investigations

## Abstract

**Objective:** To examine the timeline from submission of Coronavirus Disease 2019 (COVID)-related original articles compared with non-COVID-related original articles.

**Background:** There have been growing concerns about the speed and rigor of the review process for COVID-related articles by journals.

**Methods:** Using Dimensions, an online searchable platform, we identified PubMed-indexed journals that published >50 COVID-related articles (regardless of article type) between 1/1/2020 and 5/16/2020 and had available data on the date of article receipt. For the control group, we included consecutive full-length original investigations with available receipt date (regardless of topic) published in these journals starting from 3/1/2019 until a 1:2 ratio of COVID to non-COVID-related articles per journal was achieved.

**Results:** The final number included 294 COVID-related full-length original investigations with available article receipt dates published in 16 journals with corresponding 588 control articles from the same journals. The median time from article receipt to online publication was 20 (11-32) days for COVID-articles vs. 119 (62-182) days for controls (P<0.001). The median time to final acceptance (available for 97% of the articles) was 13 (5-23) days for COVID vs. 102 (55-161) days for controls (P<0.001). These observations were seen across all the included journals in the analysis.

**Conclusions:** In this analysis of full-length original investigations published in 16 medical journals, the median time from receipt to final acceptance of COVID-related articles was 8 times faster compared to non-COVID-related articles published in a similar time frame in the previous year. Online publication was 6 times faster for COVID-related articles compared to controls.

## INTRODUCTION

The ongoing Coronavirus Disease 2019 (COVID-19) pandemic has been the focus of an immense number of scientific publications in the recent months. While it is understandable that medical journals would strive to ensure timely publication of COVID-related articles, there have been growing concerns about the speed and rigor of the review process [1]. We aimed to quantify the timeline from submission to publication of COVID-related articles relative to non-COVID-related controls.

## METHODS

Using Dimensions, an online searchable platform that collects data on >100 million publications [2], we identified PubMed-indexed journals that published >50 COVID-related articles (regardless of article type) between 1/1/2020 and 5/16/2020. We included only journals that provided the date of article receipt. We reviewed the abstract and/or full-text of these articles so as to include-only full-length original investigations (with available receipt date). We excluded journals with fewer than 3 original articles fulfilling the above-mentioned criteria. For the control group, we included consecutive full-length original investigations with available receipt date (regardless of topic) published in these journals starting from 3/1/2019 until a 1:2 ratio of COVID to non-COVID-related articles per journal was achieved. We collected the receipt and online publication dates of included papers. We also retrieved the first authors’ revision and final acceptance dates, if reported. Median (25^th^-75^th^ percentiles) were calculated for continuous variables since their distribution was not normal. We compared the medians between COVID-related articles and controls using the Mann-Whitney U test. The statistical analyses were performed using SPSS statistical package (SPSS version 25.0, IBM Inc., Armonk, NY).

## RESULTS

Among 31 journals with >50 COVID-related articles, 20 journals provided receipt dates for published articles. Of 1,664 COVID-related articles published in these 20 journals during the study period, 299 were full-length original investigations with available article receipt dates. Four journals published <3 eligible articles (total of 5 articles in all 4), thus, were excluded. The total number of COVID-related articles included in this analysis was 294 articles from 16 journals, with corresponding 588 control articles from the same journals. The median time from article receipt to online publication was 20 (11-32) days for COVID-articles vs. 119 (62-182) days for controls (P<0.001). The median time to final acceptance (available for 97% of the articles) was 13 (5-23) days for COVID vs. 102 (55-161) days for controls (P<0.001). These observations were seen across all the included journals in the analysis (**Figure**). Among 267 COVID-related studies with available final acceptance date, 32 (12%) were accepted within 2 days of receipt. The median time to first authors’ revision (available for 58% of the articles) was 10 (5-18) vs. 71 (42-117) days for COVID and control articles (P<0.001).

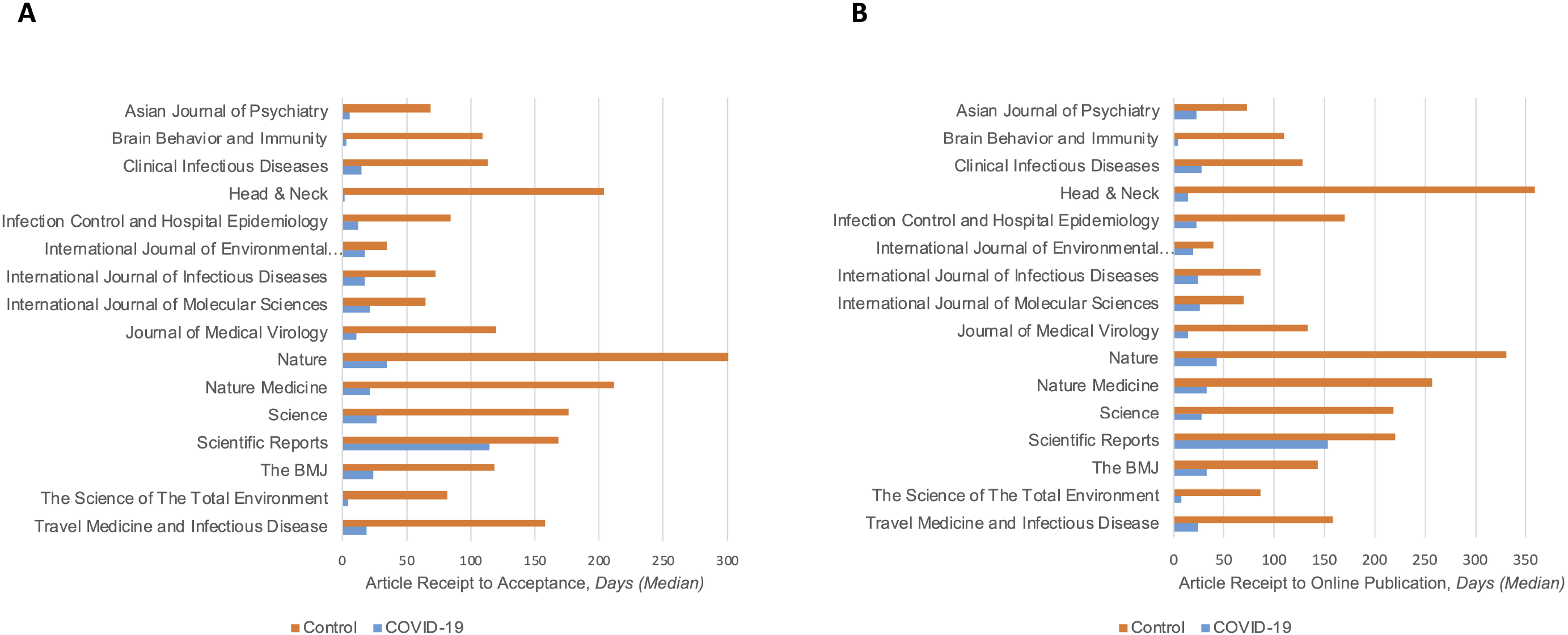
The median time from article receipt to final acceptance (Panel A) and to online publication (Panel B) for COVID-19 articles (blue) and controls (orange) in each of the included journals. The number of COVID-19 and control articles per journal, respectively, was as follows: Asian Journal of Psychiatry (7 and 14), Brain Behavior and Immunity (8 and 16), Clinical Infectious Diseases (45 and 90), Head & Neck (13 and 26), Infection Control and Hospital Epidemiology (4 and 8), International Journal of Environmental Research and Public Health (33 and 66), International Journal of Infectious Diseases (36 and 72), International Journal of Molecular Sciences (4 and 8), Journal of Medical Virology (55 and 110), Nature (8 and 16), Nature Medicine (8 and 16), Science (11 and 22), Scientific Reports (5 and 10), The BMJ (3 and 6), The Science of The Total Environment (43 and 86), and Travel Medicine and Infectious Disease (11 and 22 articles). P-value for all comparison was <0.01, except for Scientific Reports’ time to final acceptance (P=0.1) and for the BMJ’s time to final acceptance and to online publication (P=0.02 for both). COVID-19 = Coronavirus Disease 2019; BMJ = British Medical Journal

## DISCUSSION

In this case-control analysis of 882 full-length original investigations published in 16 medical journals, we found that the median time from receipt to final acceptance of COVID-related articles was 8 times faster compared to non-COVID-related articles published in a similar time frame in the previous year. Online publication was 6 times faster for COVID-related articles compared to controls. Remarkably, more than10% of COVID-related studies were accepted within 2 days of submission. While expedient dissemination of new observations is important in a pandemic due to a novel virus, this short of a timeline might adversely affect the quality of the peer review process, unintentionally spread misinformation and/or lead to retraction [3,4]. Observing a rapid rate for review of COVID science does not necessarily implicate a flawed pre-publication review process. It may be that non-COVID proceeds too slowly. At a minimum, however, we believe these observations serve as a call to journal editors to re-evaluate the timeline of handling original investigations related to COVID-19.

## Data Availability

All data are available in the manuscript

## Notes

### Competing Interest Statement

The authors have declared no competing interest.

### Funding Statement

No funding

### Author Declarations

This study used publicly available data, not related to patients, thus was exempted from IRB

